# Dual-Branch EfficientNet Architecture for ACL Tear Detection in Knee MRI

**DOI:** 10.1101/2025.09.09.25335391

**Authors:** Tharun Kota, Konstantina Garofalaki, Fiona Whitely, Elena Evdokimenko, Ellie Smartt

## Abstract

We propose a deep learning approach for detecting anterior cruciate ligament (ACL) tears from knee MRI using a dual-branch convolutional architecture. The model independently processes sagittal and coronal MRI sequences using EfficientNet-B2 backbones with spatial attention modules, followed by a late fusion classifier for binary prediction. MRI volumes are standardized to a fixed number of slices, and domain-specific normalization and data augmentation are applied to enhance model robustness.

Trained on a stratified 80/20 split of the MRNet dataset, our best model—using the Adam optimizer and a learning rate of 1e-4—achieved a validation AUC of 0.98 and a test AUC of 0.93. These results show strong predictive performance while maintaining computational efficiency. This work demonstrates that accurate diagnosis is achievable using only two anatomical planes and sets the stage for further improvements through architectural enhancements and broader data integration.

## 1. Introduction

ACL (Anterior Cruciate Ligament) is a key ligament to the stabilisation of the knee joint. Knee injuries and ACL tears are very common types of injury, especially in female athletes (Partezani Helito et al., 2023). On top of that, early diagnosis and intervention are crucial to reduce the long-term effects of the patient due to the ACL tear. Knee injuries like ACL tears are linked with lifelong effects like early onset of osteoarthritis (Partezani Helito et al., 2023). Accurate diagnosis of the injury influences the patient’s medical team whether the patient just requires rehab or will need surgical intervention. The gold standard method of diagnosis of a possible soft tissue knee injury is an MRI knee scan according to the RCR guidelines (Royal College of Radiologists, 2025). An arthroscopy procedure is the most accurate method of diagnosis, but MRI is non-invasive and prevents the patient from needing to have a surgical procedure (Royal College of Radiologists, 2025).

As with everything in health care, there are massive workloads and a shortage of staff, leading to backlogs in the Radiology departments, and anything that can reduce that workload is welcomed. This is why the goal of this project is to develop an AI model that can accurately identify ACL tears in Knee MRI. ACL tears can be easily identified to a trained eye, and the creation of this model will allow for Radiologists to focus on the more complex cases that need more nuance and experience that a lot of AI models can’t perform satisfactorily yet. A good AI can help with workload management by flagging abnormal scans, leading to faster diagnosis for the patient to get the necessary care quicker.

Current State-of-the-art approaches typically leverage 2D convolutional neural networks (CNNs) such as ResNet, EfficientNet, and DenseNet, applied slice-wise, followed by temporal pooling to aggregate predictions across slices (Raghu et al., 2019)(Liu et al., 2018). Others employ 3D CNNs to exploit volumetric information, but at the cost of higher computational resources (Xiang et al., 2025). To enhance spatial reasoning, many architectures integrate multi-branch networks, where each branch processes a different imaging plane, and the outputs are fused at a later stage [4]. Attention modules, particularly spatial attention and channel-wise attention, have shown to improve performance by focusing the model on relevant anatomical structures (Woo et al., 2018).

Despite the performance gains, several challenges remain unresolved. First, most public datasets provide only exam-level labels, lacking fine-grained annotations at the slice or voxel level. This limits the application of fully supervised learning and restricts model interpretability. As a result, recent work has explored weak supervision, multiple instance learning (MIL), and semi-supervised approaches to bridge this gap (Bai et al., 2017)(Ilse et al., 2018). Second, existing models often demonstrate poor generalization across clinical settings due to differences in MRI scanners, patient demographics, and acquisition protocols. Addressing this requires domain adaptation and multi-institutional training strategies (Ghafoorian et al., 2017).

MRI knee and other MSK (musculoskeletal) imaging is a great starting point for developing AI models, as it reduces risk to patients if the accuracy is low. This is a great first step in the development of AI algorithms for different injuries and diseases.

In this work, we trained the model with only two sequences out of the MRI knee exam – the Coronal and Sagittal planes. This was due to limited time and limited GPU space. These planes were chosen as they allow for the best visualisation of the ACL in the knee compared to the axial, which has the comparatively the worst view of the ligament. If we can confidently identify an injury with a high rate of accuracy in the test data, it could lead to advancements in the future, of fewer sequences needed to be performed in MRI departments for a diagnosis. This, in turn, will allow quicker appointments and more patients to get scanned.

## 2. MRNet Dataset

For this study, we used the Stanford MRNet dataset, which is publicly available. The dataset comprises MRI knee examinations that had 3 different sequences with different planes and scan parameters in each. The three different scan types were Sagittal T2 with fat saturation, Coronal T1 weighted, and Axial PD (proton density) with fat saturation. The different planes and scan parameters help highlight the different parts of the patient’s anatomy and can demonstrate different abnormalities all of which help with the possible diagnosis. The MRNet dataset originally included 1,370 studies, but the 120-study test set was not publicly released, leaving 1,250 labeled studies available. Labels were based on radiologist reports, primarily indicating pain, injury, or preoperative evaluation (Stanford ML Group, 2025).

The MRNet dataset was chosen due to the fact it is a benchmark dataset. A benchmark dataset is “a well-curated collection of expert-labelled data that represents the entire spectrum of disease of interest and reflects diversity of the targeted population and variation in data collection systems and methods” (Partezani Helito et al., 2023). This means that the format of the images provided was 3D arrays (no. of slices x height x width). The imaging was converted from DICOM file format, which is the ISO gold standard for medical imaging, to .npy format for ease of transfer.

Since Stanford did not release the official test set, we created our own by splitting the original training set into 80% for training and 20% for testing. To ensure balanced representation, we used a stratified split that maintains the ratio of healthy and abnormal cases. While cross-validation was considered, it was avoided due to computational and time constraints.

We leveraged a standardized MRNet data loading pipeline, which formats knee MRI studies into 3D NumPy arrays organized by imaging plane and diagnostic category. Each study was preprocessed to conform to a fixed slice count by cropping or zero-padding to the dataset’s median depth. Individual slices were normalized, resized, and converted to grayscale before being aggregated into tensors of shape. Diagnostic labels were matched using predefined CSV metadata splits. This preprocessing scheme ensures consistency across samples and facilitates efficient model training while preserving the spatial context of MRI volumes, in line with established protocols for clinical imaging tasks (Bien et al., 2018).

## 3. Model

### 3.1 Model backbone

The model backbone is EfficientNet-B2. The family of EfficientNet models was introduced by Tan and Le (Tan and Le, 2020) and was shown to achieve state-of-the-art accuracy with significantly fewer parameters and FLOPs through compound scaling of depth, width, and resolution. We chose the B2 variant as a compromise between performance and computational cost. EfficientNet-B2 achieves Top-1 and Top-5 accuracy on ImageNet comparable to Inception-v4 (Szegedy et al., 2016) and Inception-ResNet-v2 (Szegedy et al., 2017), while using approximately 5× fewer parameters and 13× fewer FLOPs. EfficientNet backbones have shown strong performance in prior medical imaging tasks, such as brain tumor classification on MRI (Ishaq et al., 2025) and COVID-19 detection in chest CT scans (Kurt et al., 2023).

We used transfer learning by initializing each EfficientNet-B2 with pre-trained ImageNet weights. This enabled faster training and convergence, and better generalization.

### 3.2 Two-branched architecture

Our model consists of a dual-branch architecture, where each branch processes slices from a different anatomical plane: sagittal or coronal. Each branch uses an EfficientNet-B2 feature extractor. Slices from each plane are processed independently by their respective EfficientNet backbone. After spatial attention (see Section 3.4) and global average pooling, slice-level features are aggregated across the temporal (slice) dimension using mean pooling. This results in one fixed-length feature vector per plane, per patient.

Each feature vector is passed through a plane-specific classifier. The classifier consists of a 30% dropout layer and a linear, fully connected (FC) layer, which produces a scalar logit. These two logits - one from the sagittal branch and one from the coronal branch - are then concatenated and passed through a final fusion classifier. The fusion classifier is also implemented as an FC layer and generates the final scalar logit, which is used for binary classification.

**Figure 1:**
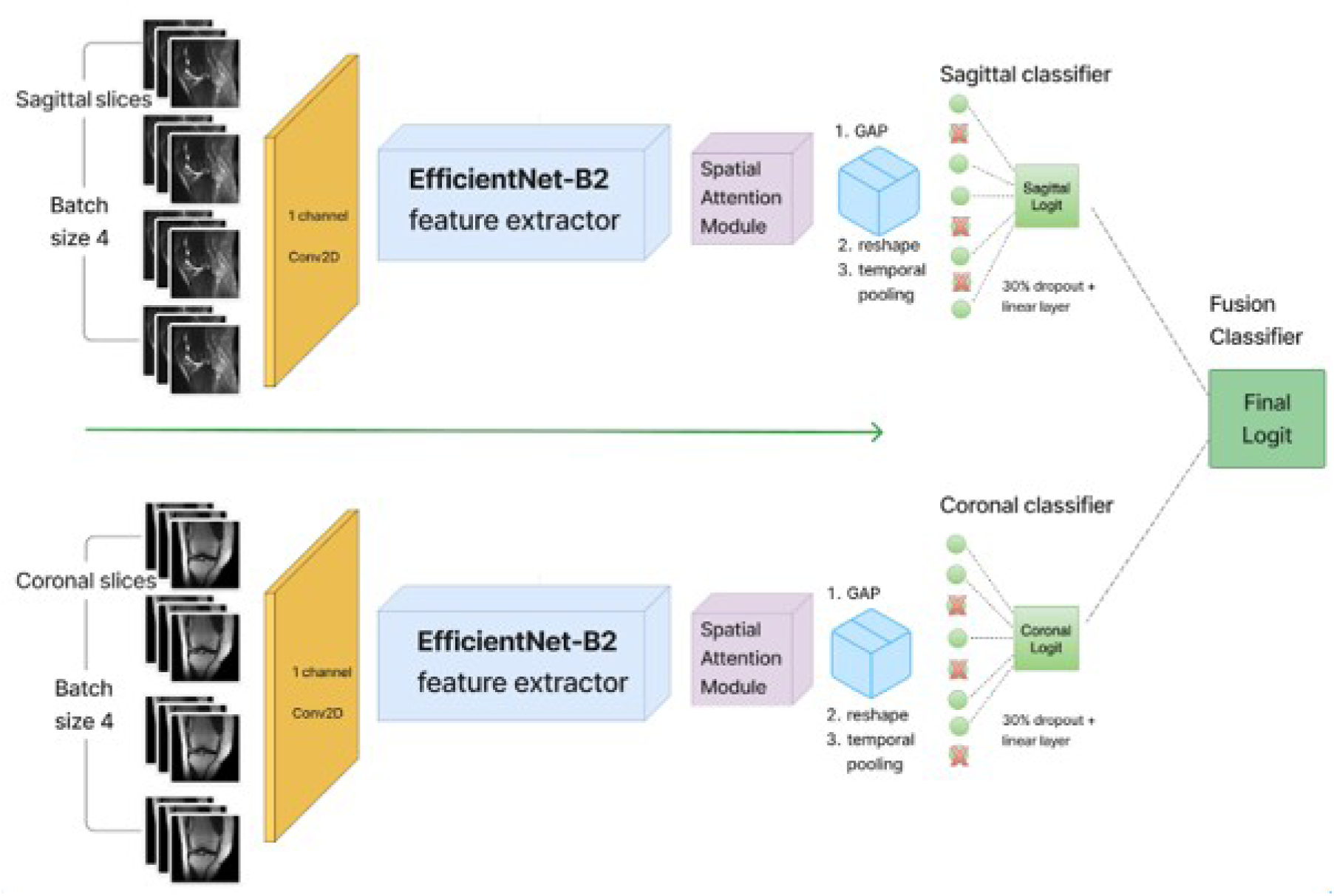
Architecture of the two-branch model. Sagittal and coronal MRI slices are independently processed through EfficientNet-B2 feature extractors and spatial attention modules. Features are pooled and classified separately, then fused by a final linear classifier.

### 3.3 Optimization for greyscale input

The first layer of the EfficientNet models is a convolutional layer that extracts low-level features such as edges and textures. It accepts A PIL image and expects a 3-channel input. Since MRI scans in our dataset are single-channel grayscale images, we modified the first convolutional layer of each EfficientNet-B2 branch to accept one channel instead of three. This was done by replacing the pretrained RGB conv layer with a new single-channel version and initializing its weights by averaging the original RGB weights across the channel dimension.

### 3.4 Spatial Attention Module

Spatial Attention Module (SAM) was employed in each of the two branches after feature extraction from EfficientNet and before global average pooling. The SAM computes a soft spatial mask from the average-pooling and max-pooling maps across the channel dimension. The mask is then applied element-wise to the features in the model (by multiplication), which enhances spatially important (e.g., lesions, anatomical boundaries) and suppresses less relevant regions of the image.

### 3.5 Implementation details

The model is implemented using PyTorch. We define a SpatialAttentionModule class, where the constructor initializes a convolutional layer to process the concatenated max- and average-pooled feature maps. We then apply a sigmoid activation. In the forward method, we produce a spatial attention map: first, we do average and max pooling across the channel, then pass results into the convolutional layer with sigmoid activation.

We define the main model class, TwoPlanesNet. It has a constructor where we initialize two EfficientNet-B2 feature extractors (for sagittal and coronal slices) and modify parameters of the first convolutional layer to accept single-channel input. The method also instantiates a SpatialAttentionModule for each branch and defines separate classifiers for each anatomical plane. We declare a final fusion classifier that takes in the concatenated outputs of the plane-specific classifiers.

In the forward method, input tensors of shape [B, S, 1, H, W] (batch, slices, channel, height, width) are reshaped, features are extracted using the model extractors that we defined in the method, then spatial attention masks are applied to the extracted features. The method also does pooling and passes features through the classifiers.

## 4. Training

### 4.1 Data Augmentation

#### 4.1.1 Normalisation

We calculated normalisation parameters using the mean and standard deviation from our own MRI images, instead of using ImageNet statistics that are commonly used in transfer learning. This approach ensures that our medical images are properly normalised according to their specific characteristics, which can be different from natural images due to MRI’s non-standardised intensity scale and different hardware characteristics.(Schwarzhans et al., 2025)

#### 4.1.2 Volume Standardisation

Since MRI volumes for different patients contain varying numbers of slices, and our batch size of 4 requires tensors with the same number of slices per plane across patients, we standardised the volume dimensions by cropping and padding. We calculated the median number of slices across both sagittal and coronal planes from the training dataset, which was 30 slices for both planes. The minimum number of slices was 17, and the maximum was 51 for the sagittal plane, while the coronal plane had a maximum of 58 slices. Volumes were cropped to this median size by taking the first N slices, where N is the median, while shorter volumes were padded with zeros. While this approach ensures consistent tensor dimensions for batch processing, future implementations would benefit from centred cropping to avoid losing important medical information from the cropped portions of the volumes.

#### 4.1.3 Augmentation

Several data augmentation techniques were used to improve model performance and prevent overfitting. Random rotation randomly rotates images by a small angle to simulate different patient positioning during MRI scans. This helps the model handle slight differences in rotation that occur during imaging. Random affine transformations apply random translation, shifting the image position, scaling, resizing the image, and shearing, shifting one part of the image parallel to a given axis, to account for variations in patient positioning and scanner settings that happen across different imaging protocols.

Random horizontal flip randomly mirrors images horizontally(Cossio, 2023). This works well since knee anatomy is symmetrical and injuries can occur on either side with similar appearance, so flipping images will still produce realistic images while effectively doubling our training data. Colour jitter randomly adjusts the brightness, contrast, and intensity of images to handle intensity variations that can occur due to different scanner settings and equipment. This makes the model more robust when applied to images from different hospitals or scanners, which is important for real-world use(Garcea et al., 2023).

### 4.2 Hyperparameters & Model Optimisation

During hyperparameter testing, all versions of the model were trained for up to 50 epochs with early stopping applied if the validation AUC did not improve for 10 consecutive epochs. The batch size was also kept constant at 4, in order to improve the generalisation from the initial batch size of 1, however, still keeping memory constraints in account since the data being processed is very large(Isensee et al., 2018). Hyperparameter tuning was done based on the optimisers and learning rate used during the training process.

#### 4.2.1 Optimisers

There were 3 different optimisers compared, namely: Adam, AdamW, and Stochastic Gradient Descent (SGD). All of these optimisers are commonly used in image classification tasks and were therefore chosen to compare their performance. The Adam optimiser was part of the original model design, so it was kept to compare its performance under different conditions. The AdamW is a version of the Adam optimiser, however, weight decay and loss-based gradient updates are decoupled, which tends to improve generalisation and overall performance in image classification, especially when paired with a learning rate scheduler, like in this case(Loshchilov and Hutter, 2017). Lastly, the performance of an SGD was added to the comparison, since it is one of the most standard optimisers used in neural network training, and it was paired with a momentum of 0.9, which is an industry standard value(Mortazi et al., 2023).

#### 4.2.2 Learning rates

The initial model applied a learning rate of 1e-5, as well as a scheduler that decreased the learning rate by 30% if it did not see an improvement of more than 1e-4 on the Binary Cross Entropy with Logits Loss criterion for more than 4 epochs. Since the scheduler already applied a learning rate decrease throughout the training process, it was decided to add one more learning rate during the hyperparameter testing phase of 1e-4, to see how starting at a higher value impacted training and performance.

## 5. Evaluation

To identify the optimal configuration, the model was trained and validated using different combinations of optimizers (Adam, AdamW, SGD) and learning rates (1e-4, 1e-5). The performance metrics for each configuration are shown in Table 1, and validation AUCs are compared in Figure 2. Detailed AUC plots for these experiments are provided in **Appendix A.**

**Figure 2:**
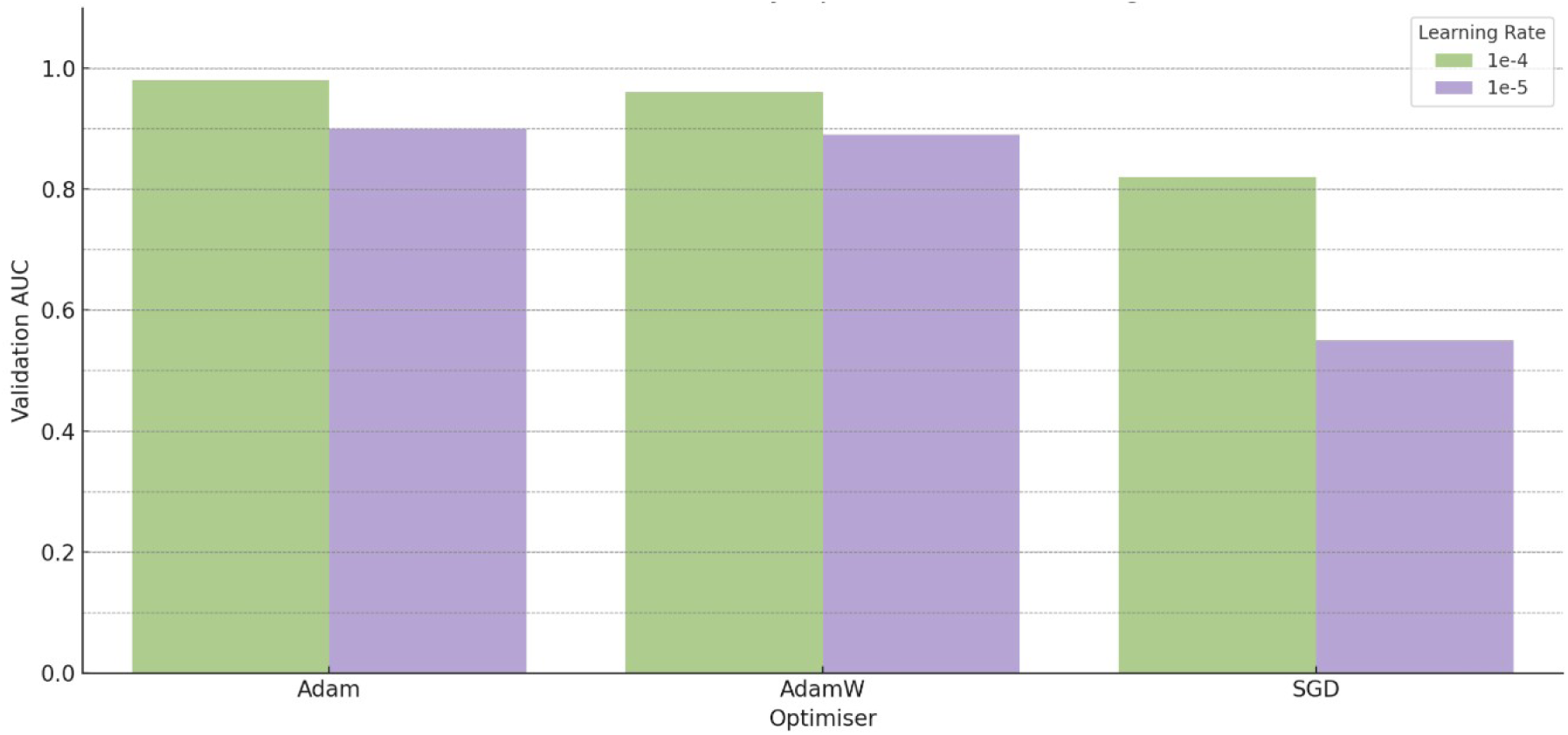
Model Validation AUC by Optimiser and Learning Rate.

**Table 1:**
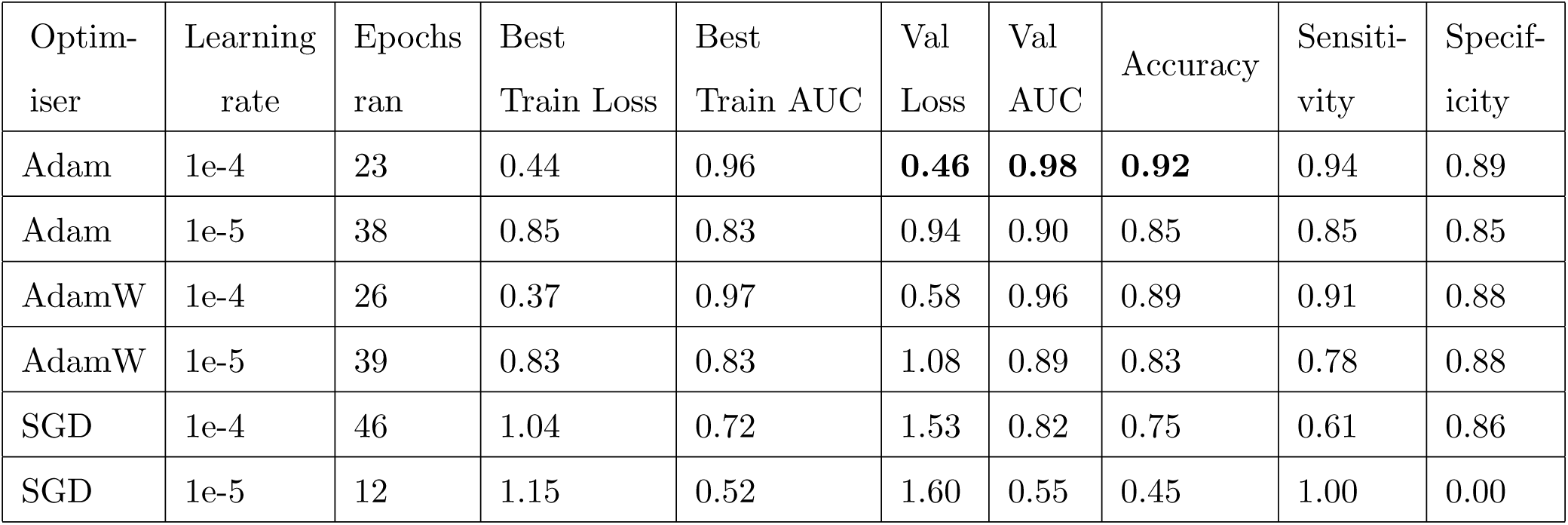
Hyperparameter tuning results.

| Optimiser | Learning rate | Epochs ran | Best Train Loss | Best Train AUC | Val Loss | Val AUC | Accuracy | Sensitivity | Specificity |
| --- | --- | --- | --- | --- | --- | --- | --- | --- | --- |
| Adam | 1e-4 | 23 | 0.44 | 0.96 | **0.46** | **0.98** | **0.92** | 0.94 | 0.89 |
| Adam | 1e-5 | 38 | 0.85 | 0.83 | 0.94 | 0.90 | 0.85 | 0.85 | 0.85 |
| AdamW | 1e-4 | 26 | 0.37 | 0.97 | 0.58 | 0.96 | 0.89 | 0.91 | 0.88 |
| AdamW | 1e-5 | 39 | 0.83 | 0.83 | 1.08 | 0.89 | 0.83 | 0.78 | 0.88 |
| SGD | 1e-4 | 46 | 1.04 | 0.72 | 1.53 | 0.82 | 0.75 | 0.61 | 0.86 |
| SGD | 1e-5 | 12 | 1.15 | 0.52 | 1.60 | 0.55 | 0.45 | 1.00 | 0.00 |

The best validation AUC and accuracy were yielded using the Adam optimiser and starting with a learning rate of 1e-4. This configuration only needed to be trained for 23 epochs until its performance plateaued at a validation loss of 0.46, a validation AUC of 0.98, and 92% accuracy. The next best performing hyperparameters combined the AdamW optimiser with a starting rate of 1e-4 and achieved a 0.58 validation loss, 0.96 validation AUC, and 89% accuracy. Overall, the Adam optimiser outperformed all other optimisers and was closely followed by the AdamW, while the SGD’s performance was significantly lower than the rest. The learning rate 1e-4 also outperformed the original one of 1e-5 when combined with all three optimisers.

The best model was further investigated by looking at performance throughout the training process.

As can be seen in Figure 3, the model’s train and validation AUCs start to stabilise after around 7 epochs of training and almost fully plateau after 15 epochs. When plotting the model’s losses over epochs, it can be seen that train loss steadily decreases with more training, but validation loss decreases during the first couple of epochs, but spikes around the 10 epochs mark, and then a slight but steady increase can be seen for the rest of the training. This behaviour can be due to slight overfitting of the training data, which hinders validation performance but does not affect the overall model’s AUC score.

**Figure 3:**
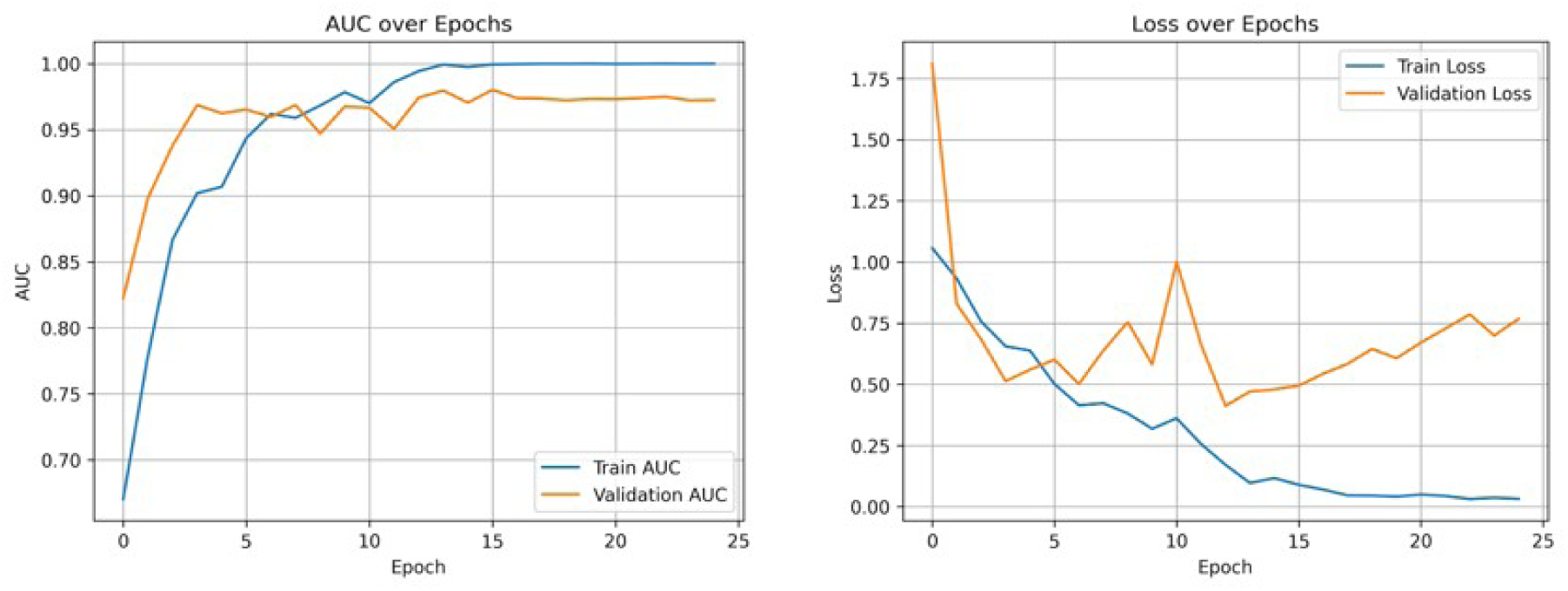
Best hyperparameters for training performance over epochs.

## 6. Results and Discussion

After training and hyperparameter tuning, the best-performing model was evaluated on the unseen test set. It achieved an AUC of 0.93, an accuracy of 89%, a sensitivity of 0.83, and a specificity of 0.91. Figure 4 summarizes the model’s performance on the test set.

**Figure 4:**
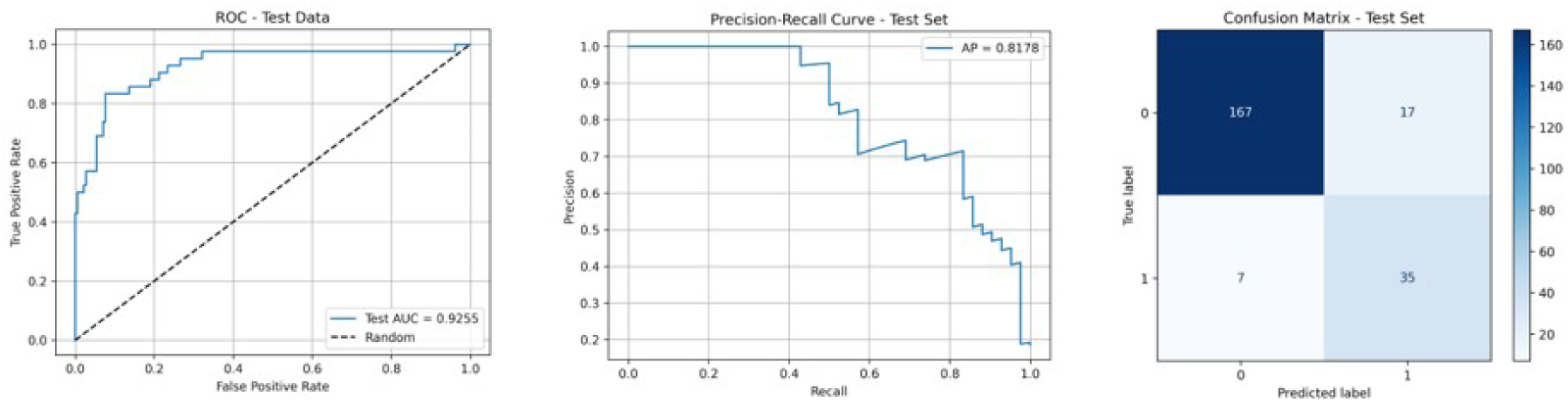
Test set performance of the best fine-tuned model

While the model showed high accuracy and AUC on the validation set, performance dropped slightly on the test set. This gap is likely due to mild overfitting during training, limited variability in the data, and the model being tuned on the validation set. Additionally, cropping/padding volumes to a fixed slice count may have discarded clinically important information. The test set, containing previously unseen samples, likely included harder or less typical cases, highlighting the need for better generalization strategies in future iterations.

## 7. Conclusion and Future Work

To improve the current architecture, one potential enhancement is to incorporate the axial plane as a third input branch alongside sagittal and coronal views. Additionally, more sophisticated fusion strategies can be explored. Instead of the current linear classifier, a multi-layer perceptron (MLP) with two fully connected layers could be used to better integrate the features from different planes. Alternatively, feature fusion through element-wise multiplication or other aggregation techniques could be investigated. In preprocessing, we used padding or cropping to standardize MRI volumes to the median slice count (30) by retaining the first 30 slices. However, using a central crop may yield better performance by increasing the likelihood of preserving medically relevant regions. Finally, experimenting with more advanced EfficientNet variants beyond the B2 model could further enhance overall performance.

## Data Availability

All the data produced in the present work are contained in manuscript

https://aimi.stanford.edu/datasets/mrnet-knee-mris

## 8. Code availability

The code for this study is available at git-hub

### Appendix A. AUC Plots for Hyperparameter Tuning

**Figure 5:**
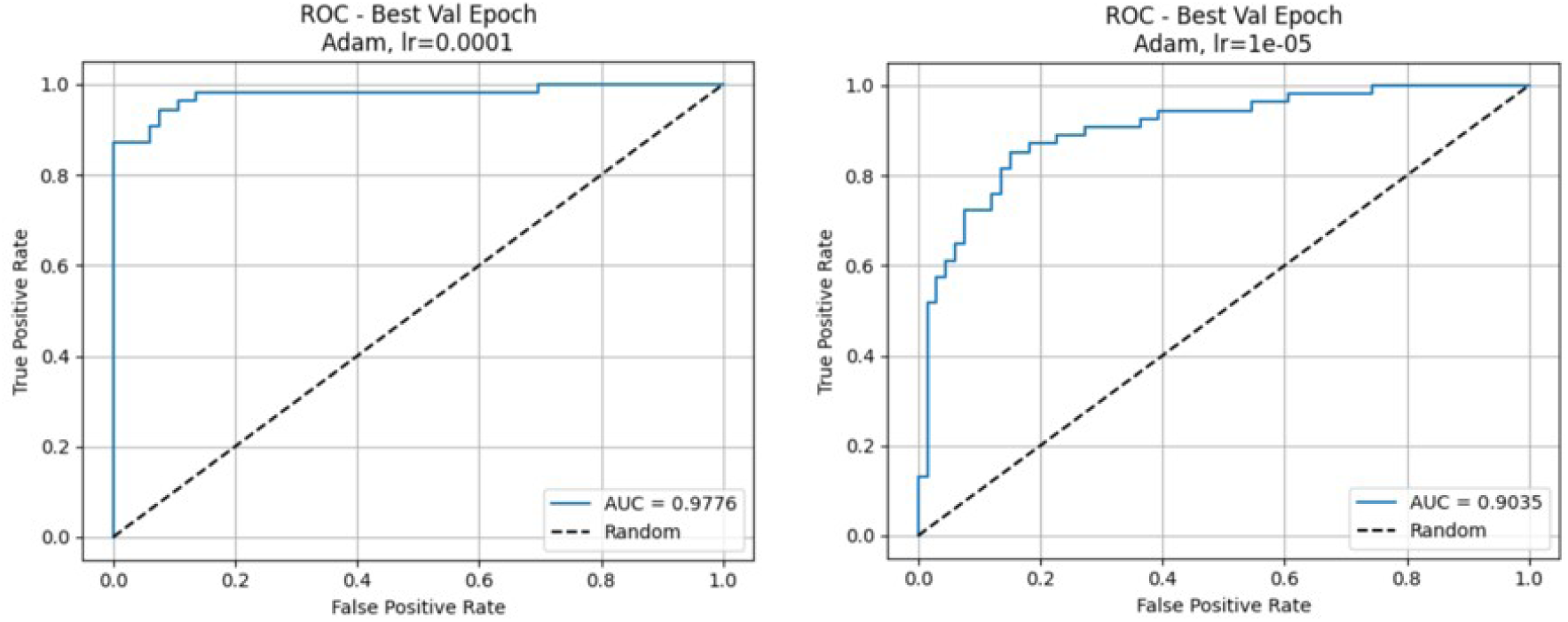
AUC comparison for Adam optimiser with starting learning rates 1e-04 and 1e-05.

**Figure 6:**
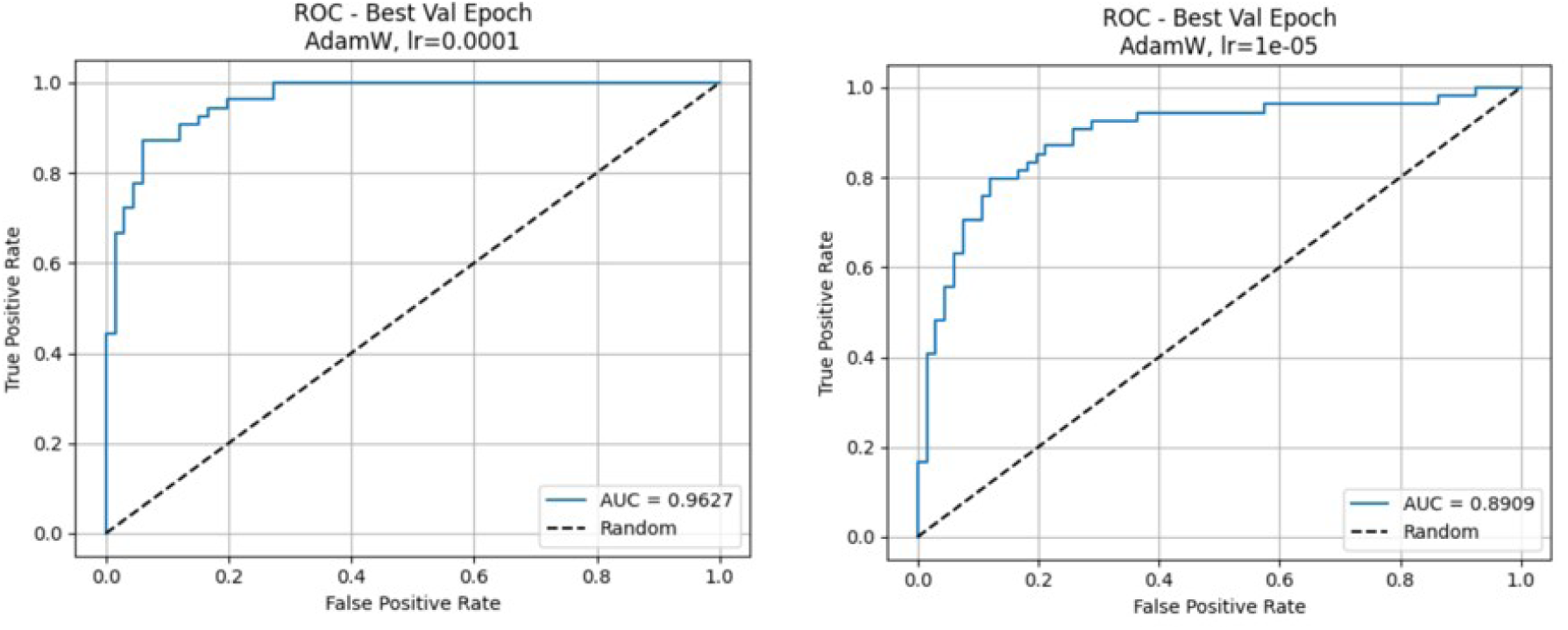
AUC comparison for AdamW optimiser with starting learning rates 1e-04 and 1e-05.

**Figure 7:**
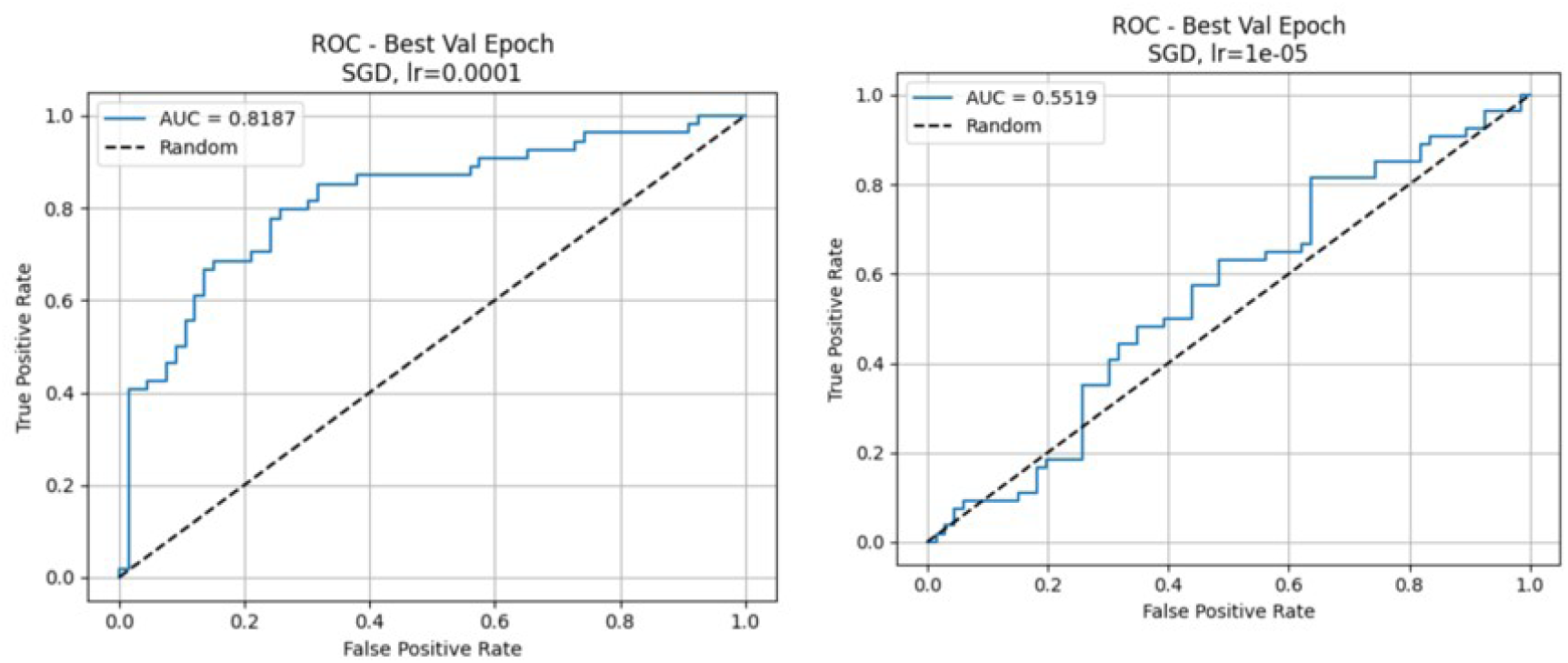
AUC comparison for SGD optimiser with starting learning rates 1e-04 and 1e-05.

### Appendix B. Performance metrics for alternative model (with two planes concatenated, and a single classifier)

In our main model, each anatomical plane (sagittal and coronal) produces its own feature vector, which is passed through a plane-specific classifier. The outputs of these classifiers are then concatenated and passed through a final fusion classifier to produce the final prediction.

As an alternative, we trained and evaluated a variant of the architecture where feature vectors from both planes were concatenated immediately and passed through a single linear classifier, bypassing the plane-specific classifiers and fusion step.

This alternative model achieved similar performance on the validation set, with AUC of 0.98 (Figure 8). However, on the unseen test set, the alternative model performance significantly dropped. The alternative model achieved AUC of 0.90 (Figure 9) on the unseen test set, while the main model AUC is 0.93 on the same unseen test set.

**Figure 8:**
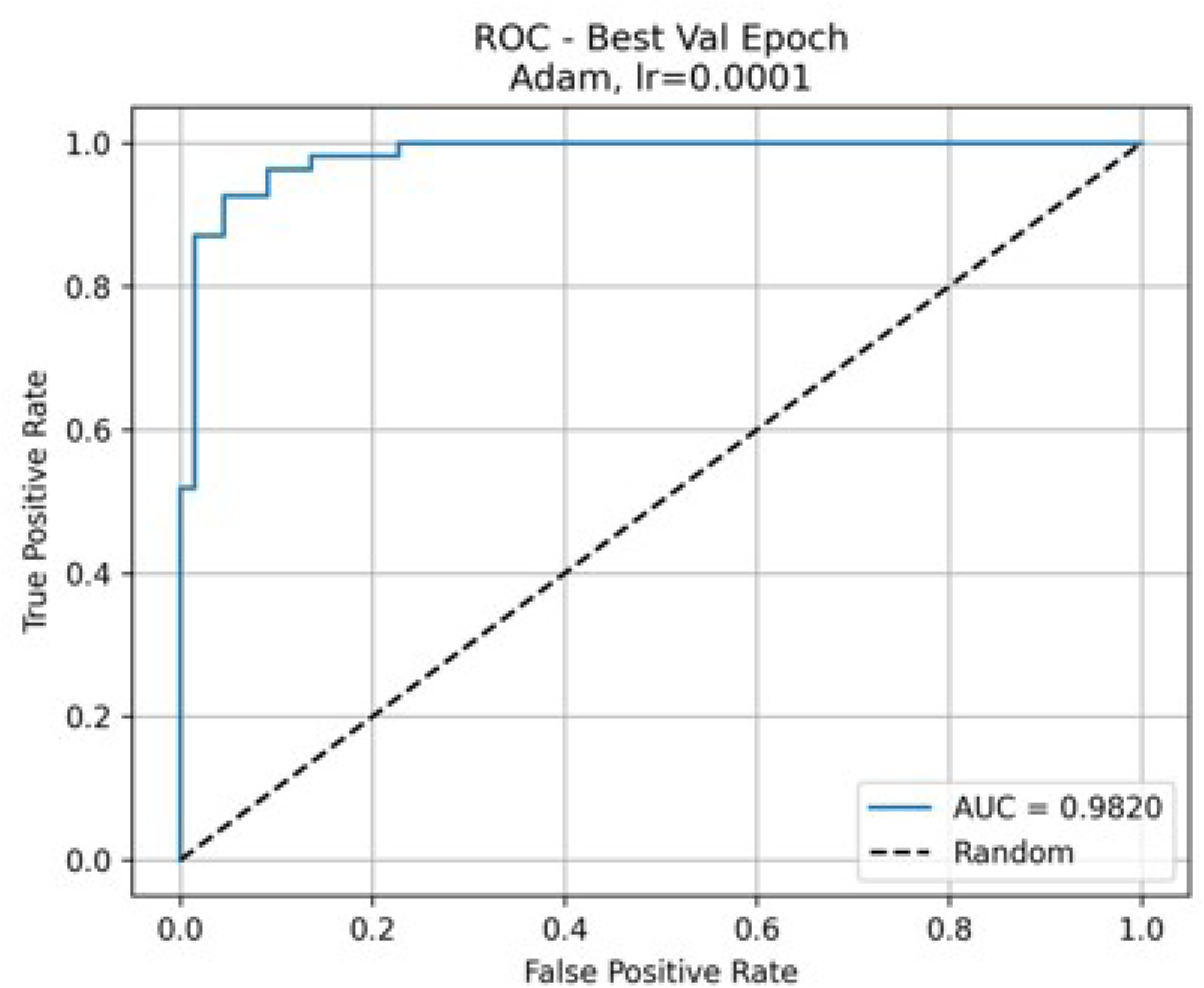
Performance metrics of the alternative model on validation set.

**Figure 9:**
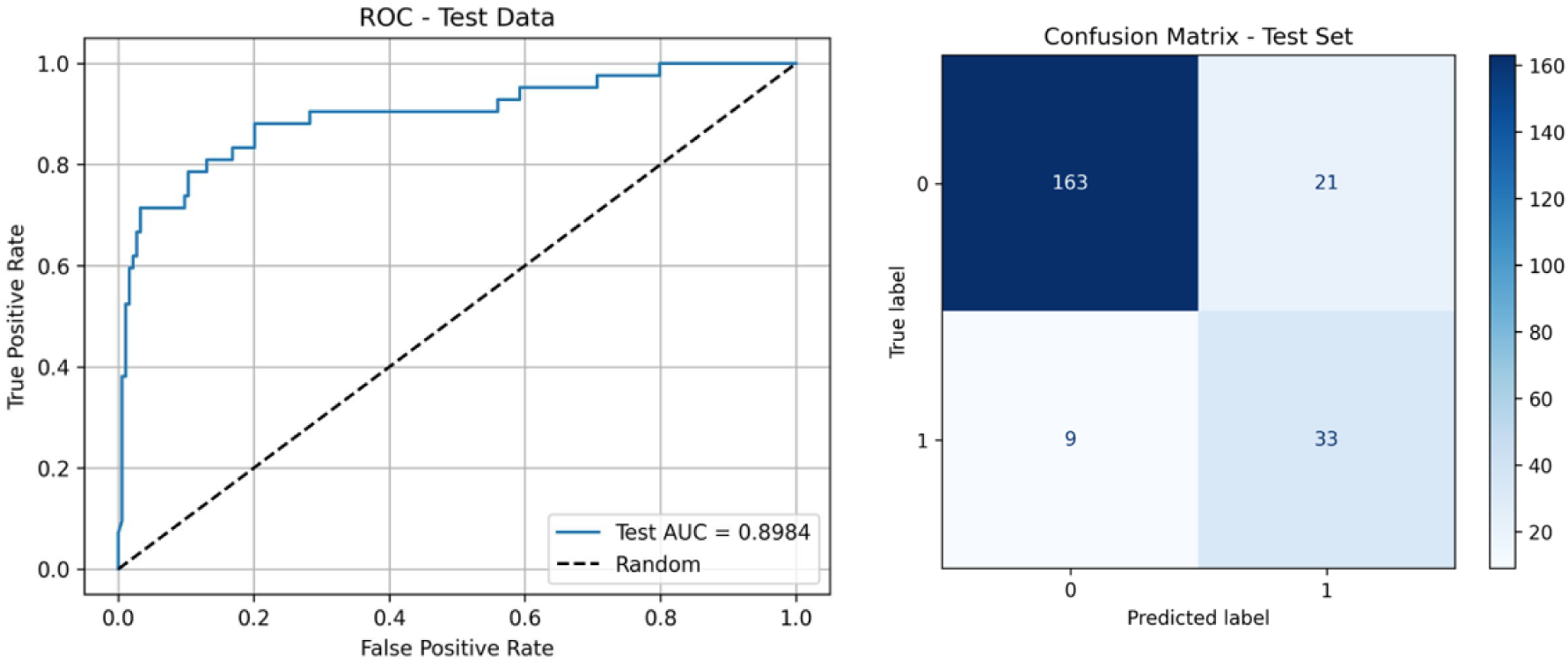
Performance metrics of the alternative model on unseen test set.

## Notes

### Competing Interest Statement

The authors have declared no competing interest.

### Funding Statement

This study did not receive any funding

